## Supplementary material for "Assessing the drivers of syphilis among men who have sex with men in Switzerland reveals a key impact of testing frequency: A modelling study"

Balakrishna *et al.*, 2020

**6 Patterns of transmission risk of syphilis** **13**

**7 Model optimization** **16**

**8 Main model fit** **18**

**9 Counterfactual scenarios** **22**

**10 Sensitivity analysis** **23**

**List of Figures** **30**

**List of Tables** **31**

**References** **32**

### 1 Model description

This section gives an overview of the model and several general assumptions. The specific details are explained in the subsequent sections. Figure 1 shows the model structure of syphilis transmission in men who have sex with men (MSM) in Switzerland. We considered the transmission of syphilis in MSM as the incidence of syphilis in MSM is much higher compared to that of heterosexuals[1]. We stratified the model by syphilis stage, syphilis detection, diagnosed HIV infection and behavioral factors to account for syphilis infectiousness, disease progression, and risk for transmission. In the main model, we used 'reported non-steady partners' (please refer section 6) as the main proxy for sexual risk. For simplicity, we assume that MSM without HIV diagnosis becomes MSM with HIV diagnosis with a constant rate of transmission as it is beyond the scope of this model and the trends in HIV incidence has remained relatively stable in last years in Switzerland.

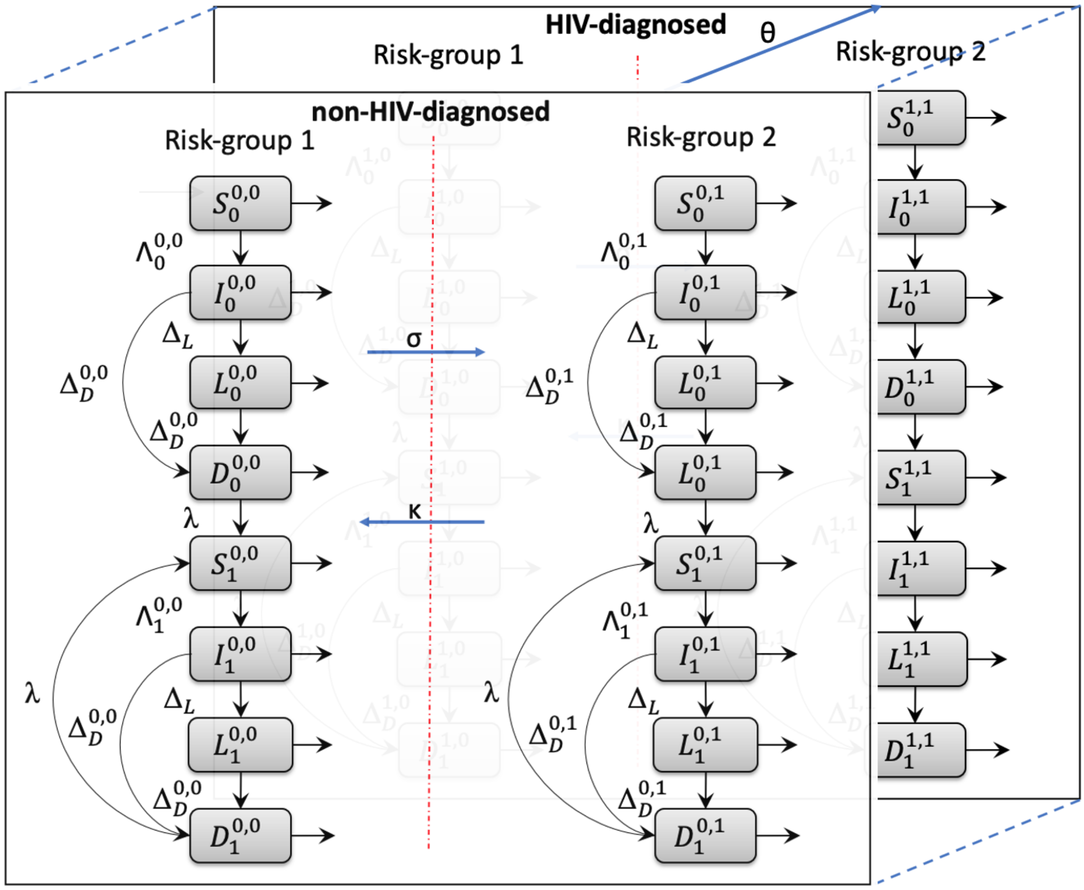

Figure 1: Model structure of syphilis transmission in men who have sex with men (MSM) in Switzerland

Note: In this document, the terms 'susceptible', 'infectious', 'non-infectious' and 'diagnosed' are all used in the context of syphilis unless explicitly mentioned.

#### 2 Model dynamics

**The dynamics of the syphilis transmission model are given by the following equations:**

$$\begin{aligned}
\dot{S}_i^{v,r} &= (M-O)\frac{S_i^{v,r}}{N} + (-1)^{1-v}\theta S_i^{1,r} + (-1)^{1-r}\sigma^v(t)S_i^{v,0} + (-1)^r\kappa^v(t)S_i^{v,1} - \Lambda_i^{v,r}S_i^{v,r} + \lambda^v i(D_0^{v,r} + D_1^{v,r}) \\
&\quad + sE \\
\dot{I}_i^{v,r} &= (M-O)\frac{I_i^{v,r}}{N} + (-1)^{1-v}\theta I_i^{1,r} + (-1)^{1-r}\sigma^v(t)I_i^{v,0} + (-1)^r\kappa^v(t)I_i^{v,1} + \Lambda_i^{v,r}S_i^{v,r} - \Delta_{IL}^{v,r}I_i^{v,r} - \Delta_{ID}^{v,r}I_i^{v,r} \\
\dot{L}_i^{v,r} &= (M-O)\frac{L_i^{v,r}}{N} + (-1)^{1-v}\theta L_i^{1,r} + (-1)^{1-r}\sigma^v(t)L_i^{v,0} + (-1)^r\kappa^v(t)L_i^{v,1} + \Delta_{IL}^{v,r}I_i^{v,r} - \Delta_{LD}^{v,r}L_i^{v,r} \\
\dot{D}_i^{v,r} &= (M-O)\frac{D_i^{v,r}}{N} + (-1)^{1-v}\theta L_i^{1,r} + (-1)^{1-r}\sigma^v(t)D_i^{v,0} + (-1)^r\kappa^v(t)D_i^{v,1} + \Delta_{ID}^{v,r}I_i^{v,r} + \Delta_{LD}^{v,r}L_i^{v,r} - \lambda^v D_i^{v,r}
\end{aligned}$$

Where,

$$\begin{aligned}
\Lambda_i^{v,r} &= \sum_{\substack{v'=\{0,1\}\\ r'=\{0,1\}}} \beta_0^{1-v}\beta_1^{v}\tau^a\eta^b(1-\eta)^{1-b}f^c(1-f)^{1-c}\left(\frac{I_0^{v',r'}+I_1^{v',r'}}{N_0^{v',r'}+N_1^{v',r'}}\right), \\
\text{if } v &= 0,\ \text{and } r=0,\ \text{and } i=0,\ \text{then } s=1,\ \text{else } s=0 \\
\text{if } r &= 1 \text{ or } r'=1,\ \text{then } a=1,\ \text{else } a=0 \\
\text{if } v &= v',\ \text{then } b=1,\ \text{else } b=0 \\
\text{if } r &= r',\ \text{then } c=1,\ \text{else } c=0
\end{aligned}$$

**Model has the following variables:**

$S_i^{v,r}$ = Susceptible; $I_i^{v,r}$ = Infected; $L_i^{v,r}$ = Non-infectious; $D_i^{v,r}$ = Diagnosed;

$v = \{0,1\}$ where, 0 = without HIV-diagnosis and 1 = with HIV-diagnosis

$r = \{0,1\}$ where, 0 = risk-group 1 and 1 = risk-group 2; stratified based on transmission risk of syphilis.

$i = \{0,1\}$ where, 0 = first episode of syphilis and 1 = recurrent episode of syphilis

$N$ = Total MSM in Switzerland; $M$ = Net-migration of MSM; $O$ = Outflow of MSM; $E$ = Inflow of MSM (See section 4)

Model parameters are given in section 3. Detailed explanation of the force of infection ($\Lambda_i^{v,r}$) is given in section 3.3

### 3 Model parameters

#### 3.1 Free parameters of the model

There are 6 free (or partly restricted) parameters in the model which are fitted in the optimization procedure (Table 1). See section for more details.

Table 1: Free parameters of the model obtained for the main model fit assuming MSM with non-steady partners as the transmission risk of syphilis

| Variable | Reason | Units | *ad-hoc* prior bounds (lower - upper) | Posterior distribution [Median (IQR)] |
|---|---|---|---|---|
| $\beta_0$ | Transmission rate of syphilis in MSM without HIV diagnosis | $\text{year}^{-1}$ | (0 - 5) | 1.36 (1.22 - 1.52) |
| $\beta_1$ | Transmission rate of syphilis in MSM with HIV diagnosis | $\text{year}^{-1}$ | (0 - 30) | 21.80 (21.68 - 21.91) |
| $\theta$ | Transmission rate of HIV | $\text{year}^{-1}$ | (0 - 0.1) | 0.0038 (0.0037 - 0.0039) |
| $f$ | Risk-sorting probability | - | (0.5 - 1) | 0.65 (0.62 - 0.67) |
| $\text{inf}_0(0)$ | Initial number of MSM without HIV diagnosis infected with syphilis in 2006 | - | (0 - 10) | 5.44 (2.32 - 7.88) |
| $\text{inf}_1(0)$ | Initial number of MSM with HIV diagnosis infected with syphilis in 2006 | - | (0 - 10) | 1.89 (1.35 - 2.55) |

IQR = Interquartile range; MSM = Men who have sex with men;

#### 3.2 Non-free parameters of the model

All other parameters than the free parameters were obtained from literature or derived from data obtained from Gay survey, Voluntary Counselling and Testing (VCT) data, and the SHCS data. (Table 2)

Table 2: Non-free parameters of the model obtained for the main model fit assuming MSM with non-steady partners as the transmission risk of syphilis

| Variable | Value | Reason | Source |
|---|---|---|---|
| $\eta^1$ | 0.34 | Serosorting probability among MSM with HIV diagnosis | Derived from VCT data (section 3.3) |
| $\eta^0$ | 0.91 | Serosorting probability among MSM without HIV diagnosis | Derived from VCT data (section 3.3) |
| $\tau$ | 3.167 | Relative risk due to reported non-steady partners | Derived from SHCS (section 3.3) |

| | | | |
|---|---|---|---|
| $\Delta_{IL}^{v,r}$ | 4 year$^{-1}$ | Rate of becoming non-infectious from infected: Obtained as the inverse of average time to become non-infectious = 3 months | STD Guidelines[2] |
| $\Delta_{LD}^{1,r}$ | 1 year$^{-1}$ | Rate of becoming diagnosed from non-infectious for MSM with HIV diagnosis | Derived (section 3.4) |
| $\Delta_{LD}^{0,r}$ | 0.5 year$^{-1}$ | Rate of becoming diagnosed from non-infectious for MSM without HIV diagnosis | Derived (section 3.4) |
| $\Delta_{ID}^{v,r}$ | 16 year$^{-1}$ | Rate of becoming diagnosed from infected | Derived (section 3.4) |
| $\lambda$ | 12 year$^{-1}$ | Rate of becoming susceptible for subsequent episode of syphilis from diagnosed: Obtained as the inverse of average time to become susceptible again after diagnosis = 1 month | STD Guidelines[2] |
| $\kappa^1$ | 0.74 year$^{-1}$ in 2006 to 0.45 year$^{-1}$ in 2017 | Rate of switching from nsP to without nsP for MSM with HIV diagnosis: Obtained by time-to-event survival analysis | Derived from the SHCS (section 6) |
| $\sigma^1$ | 0.49 year$^{-1}$ in 2006 to 0.41 year$^{-1}$ in 2017 | Rate of switching back to nsP from without nsP for MSM with HIV diagnosis: Obtained by time-to-event survival analysis | Derived from the SHCS (section 6) |
| $\kappa^0$ | 0.54 year$^{-1}$ | Rate of switching from nsP to without nsP for MSM without HIV diagnosis: Assumed to be average rate of switching from MSM with HIV diagnosis with nsP to MSM with HIV diagnosis without nsP | Model assumption (section 6) |
| $\sigma^0$ | 1.44 year$^{-1}$ in 2006 to 1.48 year$^{-1}$ in 2017 | Rate of switching back to nsP from without nsP for MSM without HIV diagnosis: Obtained using time-to-event analysis based on the $\kappa^0$ | Derived from Gay Survey (section 6) |

#### 3.3 Rate of becoming Infected from Susceptible

Susceptible MSM become infected with syphilis at rate equal to the force of infection ($\Lambda_i^{v,r}$) given by:

$$\begin{aligned}
\Lambda_i^{v,r} &= \sum_{\substack{v'=\{0,1\}\\ r'=\{0,1\}}} \beta_0^{1-v}\beta_1^{v}\tau^{a}\eta^{b}(1-\eta)^{1-b}f^{c}(1-f)^{1-c}\left(\frac{I_0^{v',r'}+I_1^{v',r'}}{N_0^{v',r'}+N_1^{v',r'}}\right),\\
\text{if r} &= \text{1 or r' = 1, then a=1, else a=0}\\
\text{if v} &= \text{v', then b=1, else b=0}\\
\text{if r} &= \text{r', then c=1, else c=0}
\end{aligned}$$

where,

$$\beta_0 = \text{transmission rate of syphilis in MSM without HIV diagnosis}$$
$$\beta_1 = \text{transmission rate of syphilis in MSM with HIV diagnosis}$$
$$\text{v} = \text{HIV-diagnosis status of self and v' = HIV-diagnosis status of the partner}$$
$$\text{r} = \text{risk-status of self and r' = risk-status of the partner}$$
$$\eta = \text{serosorting probability}$$
$$f = \text{risk-sorting probability}$$
$$I_0^{v',r'} + I_1^{v',r'} = \text{total number of infected MSM given a HIV-diagnosis status and a risk-status}$$
$$N_0^{v',r'} + N_1^{v',r'} = \text{total number of MSM given a HIV-diagnosis status and a risk-status}$$
$$\tau = \text{Relative risk due to reported non-steady partners}$$

A susceptible MSM can be infected with syphilis due to contacts with an MSM infected with syphilis. However, the possible infectious contacts are not completely random. These contacts depend on the HIV and risk status of self and the partner. In our model, we account for the contact selection through serosorting and risk-sorting probabilities. We assume that the risk-sorting and serosorting are mutually independent.

Serosorting probability ($\eta$) is defined as the probability of contact between the partners of same sero-status (HIV status). We derived the serosorting probabilities for MSM with and without HIV diagnosis using the Voluntary Counselling and Testing (VCT) data and were found to be 0.34 and 0.90 respectively. In other words, given a HIV-diagnosed MSM, the probability of contact with an HIV-diagnosed MSM is 0.34 whereas the probability of contact with a MSM without HIV diagnosis is $1 - 0.34 = 0.66$. Similarly, given a MSM without HIV diagnosis, the probability of contact with an MSM without HIV diagnosis is 0.91 whereas the probability of contact with a HIV-diagnosed MSM is $1 - 0.91 = 0.09$.

Similarly, risk-sorting probability ($f$) is defined as the probability of contacts between the partners of same risk behaviour. We assumed risk-sorting probability to be a time-independent free parameter. We further assumed the probability of contact with an MSM of risk-group A given an MSM of risk-group B to be equal to the probability of contact with an MSM of risk-group B given an MSM of risk-group A and estimated $f$ by fitting the model fit.

#### 3.4 Rate of becoming Diagnosed

MSM can be diagnosed with syphilis when they are infected with syphilis and are in either infected state (infectious) or the non-infectious state. The rates of becoming diagnosed from infected and non-infectious are given by $\Delta_{LD}$ and $\Delta_{ID}$ respectively.

80% of the syphilis cases reported in the year 2017 were found in the primary and secondary stage of syphilis (average time of 3 months). i.e., 80% of the infected MSM in the model were diagnosed with syphilis. The rest 10% of the infected MSM become non-infectious. The rate of becoming non-infectious from infected = 4 (see table 2). By Fellers theorem, the probability of the next event is proportional to the rate. This corresponds to the following equation:

$$
\begin{aligned}
\frac{\Delta_{ID}}{\Delta_{IL}} &= \frac{80}{20} \\
\text{or, } \Delta_{ID} &= 4\Delta_{IL} = 4 * 4 \\
\Delta_{ID} &= 16 \\
\text{where,} & \\
\Delta_{ID} &= \text{Rate of becoming diagnosed from infected} \\
\Delta_{IL} &= \text{Rate of becoming non-infectious from infected} = 4
\end{aligned}
$$

The MSM in the non-infectious compartment will be diagnosed with syphilis depending on the routine screening rates. In the Swiss HIV Cohort Study (SHCS), MSM are routinely screened for syphilis once a year. Hence, We assume that the average time to routine screening for MSM with HIV diagnosis to be 1 year. Although, the recommendation of syphilis screening in MSM without HIV diagnosis is once per year, not everyone is routinely tested every year. Based on the European Men-Who-Have-Sex-With-Men Internet Survey (EMIS-2017), only 50% of the participants had syphilis screening in the past year[3, 4]. Hence, we assumed the average time to diagnosis in MSM without HIV diagnosis to be 2 years. The corresponding rates of switching from non-infectious to diagnosed for MSM with and without HIV diagnosis are 1 and 0.5 $year^{-1}$ respectively.

$$
\begin{aligned}
\text{i.e., } \Delta_{LD}^{1,r} &= 1 \\
\Delta_{LD}^{0,r} &= 0.5 \\
\text{where,} & \\
\Delta_{LD}^{1,r} &= \text{Rate of becoming diagnosed from non-infectious for MSM with HIV diagnosis} \\
\Delta_{LD}^{0,r} &= \text{Rate of becoming diagnosed from non-infectious for MSM without HIV diagnosis}
\end{aligned}
$$

### 4 Demographic dynamics

According to the Federal Statistical office (BAG), the total population of Switzerland in 2012 was 7.95 million[5] and the estimated number of MSM in Switzerland between the age group of 15 to 64 in 2012 was 80000[6]. Hence, 1.01% of the total population of Switzerland corresponds to MSM between the age group of 15 to 64 in 2012. By assuming that the percentage of MSM in Switzerland to be constant (1.01%) over the time period 2006 to 2019, we estimate the demographic dynamics of MSM in each year based on the demographic dynamics of Switzerland. (Figure 2)

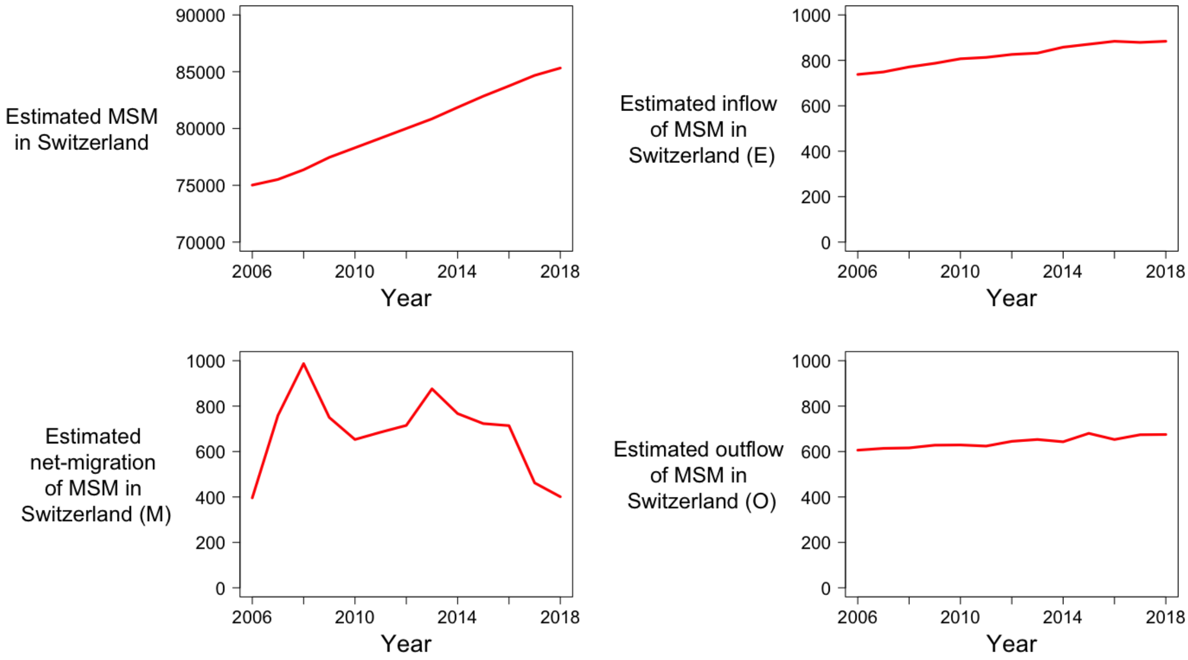

Figure 2: Estimated demographic dynamics of MSM in Switzerland

We assume that the MSM between the age group of 15 to 64 are sexually active. We further make a simplifying assumption that the the number MSM who become sexually active and are of low risk and are never infected with syphilis or HIV in a given year corresponds to 1.01% of the new births in Switzerland in the respective year. In other words, this corresponds to the inflow of MSM ($E$) into the model who enter as MSM without HIV diagnosis without nsP who never had syphilis.

We assume that the number of MSM who either die or become sexually inactive in a given year corresponds to 1.01% of the new deaths in Switzerland the respective year and the deaths occurs at a same rate in all the compartments irrespective of HIV and syphilis infection. In other words, this corresponds to the outflow of MSM ($O$) that is proportional to the size of the compartments.

We assume the number of sexually active MSM entering Switzerland in a given year corresponds to 1.01% of the net-migration (difference of immigration and emigration) in Switzerland in the respective year. We further assume that the proportion of MSM in different compartments in Switzerland is equal to the proportion of MSM in different compartments of other countries from which the MSM are immigrated to

Switzerland i.e., distribution of MSM immigrants across different compartments is equal to the distribution of MSM emigrants across different compartments. In other words, this corresponds to the net-immigration of MSM ($M$) that is proportional to the size of the compartments.

We make these simplifying assumptions as we do not observe the demographic dynamics of MSM in Switzerland directly and these assumptions allow us to link the demographic dynamics of MSM to the demographic dynamics of Switzerland.

### 5 Initial conditions

The model runs from the years 2006 to 2018. Hence, the initial population of a compartment corresponds to the value of respective compartment as of 01-01-2006.

The initial conditions depend on the definition of the risk-group 1 and 2. (Please refer to section 6 for more details)

Risk-group 1 ($r = 0$) → MSM without non-steady partners (MSM without nsP)

Risk-group 2 ($r = 1$) → MSM with non-steady partners (MSM with nsP)

#### 5.1 Susceptible (S)

We determine the initial conditions for susceptible population in different compartments of HIV-diagnosed from the SHCS data and are given by:

$$\begin{aligned}
S_0^{1,0} &= \text{MSM with HIV diagnosis and without nsP who never had syphilis before } 01\text{-}01\text{-}2006_i^{1,r} \\
S_1^{1,0} &= \text{MSM with HIV diagnosis and without nsP who had a syphilis episode before } 01\text{-}01\text{-}2006_i^{1,r} \\
S_0^{1,1} &= \text{MSM with HIV diagnosis and with nsP who never had syphilis before } 01\text{-}01\text{-}2006_i^{1,r} \\
S_1^{1,1} &= \text{MSM with HIV diagnosis and with nsP who had a syphilis episode before } 01\text{-}01\text{-}2006_i^{1,r}
\end{aligned}$$

Due to the limited information, we assume that the initial conditions for the susceptible MSM without HIV diagnosis are given by the following equations:

$$\begin{aligned}
S_0^{0,0} &= x(1-y)(1-p) \\
S_1^{0,0} &= xy(1-p) \\
S_0^{0,1} &= x(1-y)p \\
S_1^{0,1} &= xyp
\end{aligned}$$

Where,

$x$ = Total number of MSM without HIV diagnosis in Switzerland in 2006 = Total number of MSM in Switzerland in 2006 - Total number of MSM in the SHCS

$y$ = Prevalence of syphilis among MSM without HIV diagnosis = 7% = 0.07 (Source: EMIS-2017)

$p$ = Proportion of MSM without HIV diagnosis and with nsP = 0.735 (see section 6)

#### 5.2 Infected (I)

We do not directly observe the initial number of infected individuals for both MSM with and without HIV diagnosis. As there is no information on the relative numbers of syphilis cases across compartments for MSM without HIV diagnosis, we assume equal numbers of MSM infected with syphilis in all syphilis infected compartments of MSM without HIV diagnosis. It should be noted that while such a uniform distribution may be considered unrealistic, this assumption has only a marginal effect on the model dynamics, because the distribution of initial cases only affects the initial force of infection and due to the short infectious period of syphilis (3 months), the distribution across compartments quickly reaches a state which is governed not anymore by the initial conditions but by the other model parameters. For MSM with HIV diagnosis, we assume that the relative numbers of initial syphilis cases is given by the SHCS, i.e. we assume that the initial number of MSM infected with syphilis in a HIV-diagnosed MSM compartment to be proportional to the diagnosed cases of syphilis in 2006 in the respective compartment. Based on these assumptions, the initial conditions for the infected can be given by the following equations:

$$\begin{aligned}
I_0^{0,0} &= I_1^{0,0} = I_0^{0,1} = I_1^{0,1} = \text{inf}_0(0) \\
I_0^{1,0} &= \text{inf}_1(0) \\
I_1^{1,0} &= \text{inf}_1(0)(\text{Diagnosed in } 2006_1^{1,0}/\text{Diagnosed in } 2006_0^{1,0}) \\
I_0^{1,1} &= \text{inf}_1(0)(\text{Diagnosed in } 2006_0^{1,1}/\text{Diagnosed in } 2006_0^{1,0}) \\
I_1^{1,1} &= \text{inf}_1(0)(\text{Diagnosed in } 2006_1^{1,1}/\text{Diagnosed in } 2006_0^{1,0})
\end{aligned}$$

Where,
$\text{inf}_0(0)$ and $\text{inf}_1(0)$ are free parameters obtained by the model fit.

#### 5.3 Non-infectious (L)

We do not have any information on the individuals who are non-infectious. We assume it be 0 for all compartments. These compartments are populated over the period depending on the infected compartments.

$$L_i^{v,r} = 0$$

#### 5.4 Diagnosed (D)

We assume the starting state of all diagnosed compartments to be 0. These compartments are populated over the period depending on the infected and non-infectious compartments.

$$D_i^{v,r} = 0$$

### 6 Patterns of transmission risk of syphilis

In our main model, we defined transmission risk of syphilis based on the 'MSM with non-steady partners' in the following way:

Risk-group 1 ($r = 0$) $\rightarrow$ MSM without non-steady partners (MSM without nsP)
Risk-group 2 ($r = 1$) $\rightarrow$ MSM with non-steady partners (MSM with nsP)

For MSM with HIV diagnosis, proportion of MSM with nsP was estimated directly using the SHCS data for years 2006 to 2018 (figure 3).

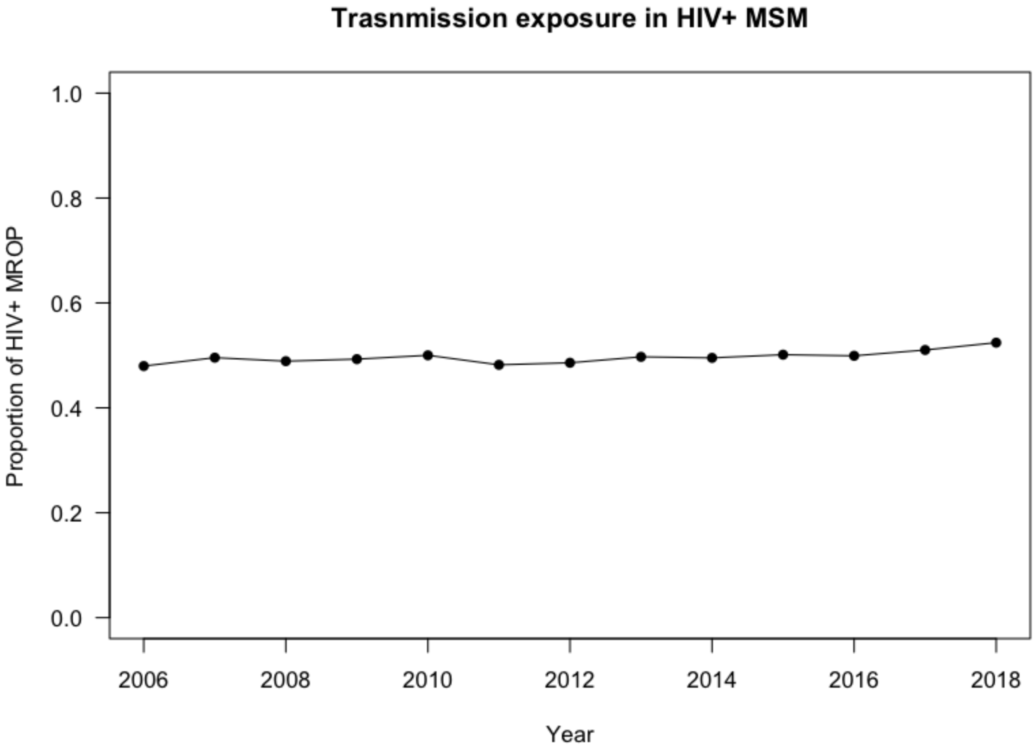

Figure 3: Proportion of MSM with HIV diagnosis and with non-steady partners

The rate of switching from MSM without nsP to MSM with nsP ($\sigma^1$) and the rate of switching from MSM with nsP to MSM without nsP ($\kappa^1$) were calculated by time-to-event analysis for each year. (figure 4)

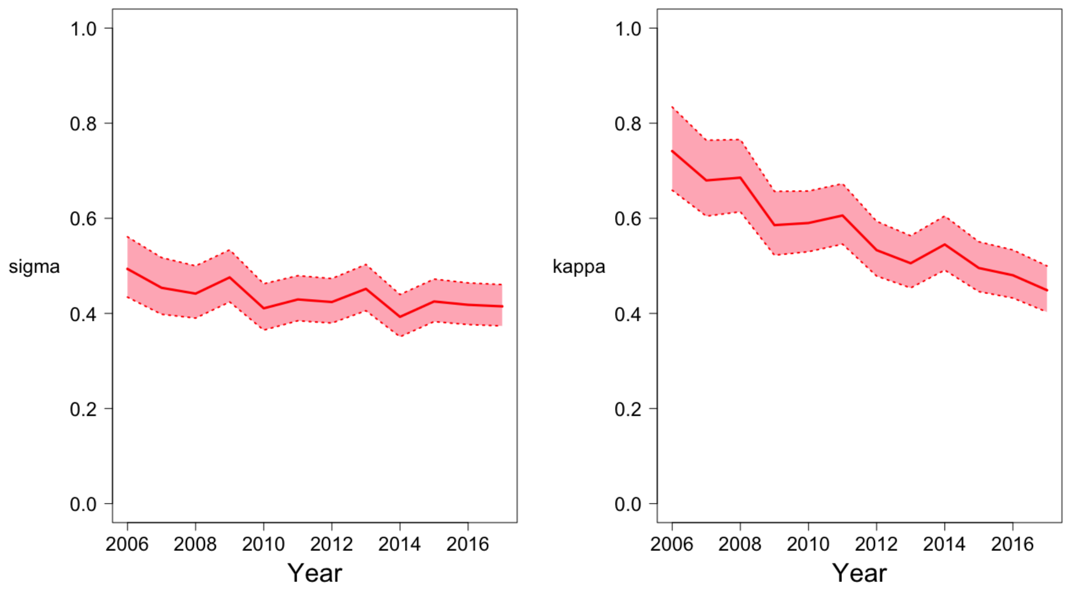

Figure 4: Estimated sigma ($\sigma^1$) and kappa ($\kappa^1$) for MSM with HIV diagnosis in Switzerland

Whereas for MSM without HIV diagnosis, proportion of MSM with and without nsP were obtained from Gay survey. However, the data is not longitudinal, and the data was available for only several years (2004, 2007, 2009, 2012 and 2014). Hence, a linear interpolation was used to estimate data points between 2004 and 2012 and constant extrapolation was used to estimate data points after 2004 as shown in figure 5.

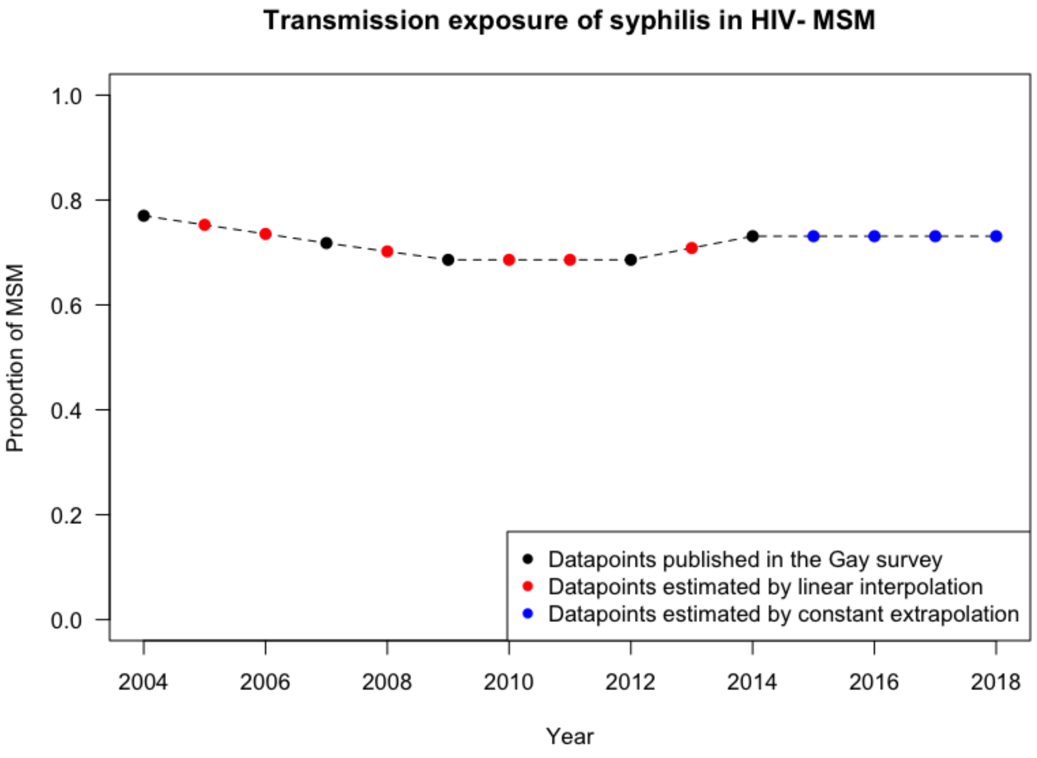

Figure 5: Proportion of MSM without HIV diagnosis and with non-steady partners

We assumed the rate of switching from MSM with nsP to MSM without nsP among MSM without HIV diagnosis to be equal to the average rate of switching from MSM with nsP to MSM without nsP among MSM with HIV diagnosis. i.e., $\kappa^0 = 0.54$ and thereby estimated the rate of switching from MSM without nsP to MSM with nsP among MSM without HIV diagnosis ($\sigma^0$). (figure 6)

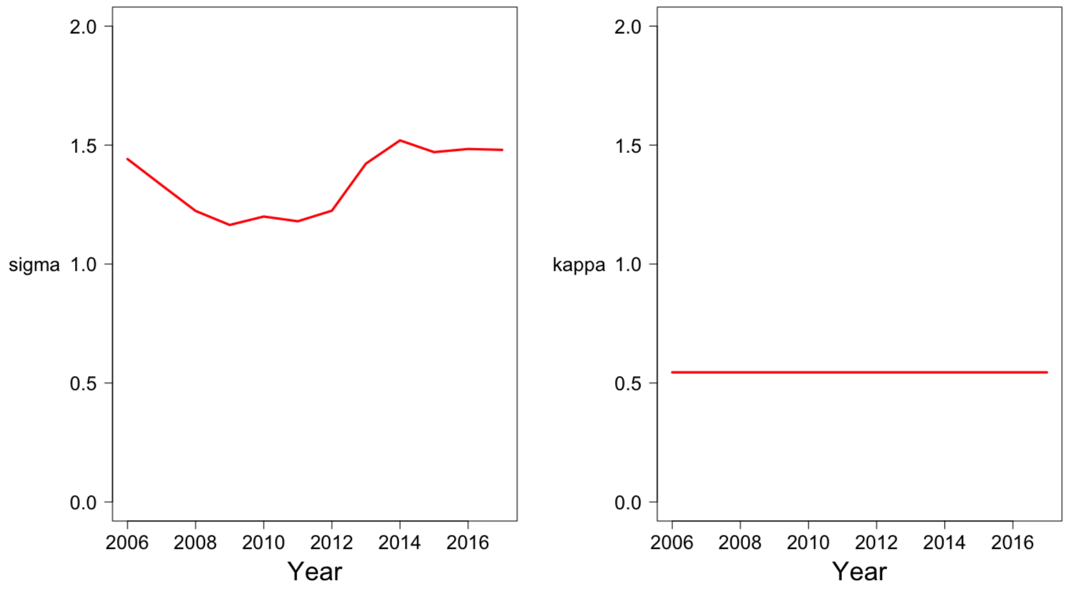

Figure 6: Estimated sigma ($\sigma^0$) and kappa ($\kappa^0$) for MSM without HIV diagnosis in Switzerland

### 7 Model optimization

The model optimization was performed by minimising the sum of squared weighted residuals between each observed and predicted datapoint. We further used simplified Markov chain Monte Carlo (MCMC), using an adaptive Metropolis algorithm and including a delayed rejection procedure, with prior values specified by the distribution of the parameters to obtain their posterior distributions. We used independent and uniform priors within *ad hoc* bounds for the free parameters.

We used the following observed datapoints for optimization of the model: estimated number of HIV-diagnosed MSM in Switzerland (section 4), estimated first/incident episode of syphilis in MSM with HIV diagnosis and without non-steady partners, estimated first/incident episode of syphilis in MSM with HIV diagnosis and with non-steady partners, subsequent episodes of syphilis in MSM with HIV diagnosis, and total cases of syphilis reported in MSM with HIV diagnosis in Switzerland from 2006 to 2017.

20 random parameter sets of 6 free parameters were generated using a latin hypercube sampling algorithm. For each of these parameter sets, models were fit to minimise the goodness of fit (sum of squared weighted residuals between the observed and predicted datapoints). Use of latin hypercube sampling fit ensures that we find a global minimum of the goodness of fit despite the randomly selected free parameters from the uniform *ad-hoc* bounds. The goodness of fit of a model was estimated using the function 'modCost' of FME package. This function is central to parameter identifiability analysis, model fitting or running a Markov chain Monte Carlo as described in [7]. We estimated the squared residual (difference between each of the observed and predicted datapoint) weighted by an error equal to square-root of predicted datapoint. Mathematically,

$$\begin{aligned}
\text{goodness of fit} &= \sum_{i=1}^{n} \left( \frac{x_i - y_i}{\sqrt{x_i}} \right)^2 \\
\text{where,} & \\
n &= \text{Number of datapoints} \\
x_i &= \text{Predicted datapoint i} \\
y_i &= \text{Observed datapoint i}
\end{aligned}$$

Model fitting was performed using the function 'modFit' to identify the parameter set which yields the best goodness of fit (lowest) using 'L-BFGS-B' method. We decided to use 'L-BFGS-B' since this method allows giving upper and lower bounds for the parameters of interest. We selected the model which yielded the best goodness of fit.

We assessed the identifiability of the free parameters using the function 'colin' of FME package. The larger the collinearity value, the less identifiable the parameter based on the data. In general a collinearity value less than about 20 is "identifiable". As shown in figure 7, the collinearity value when we consider all our 6 free parameters was 19.13. Hence, all free parameters are identifiable even when they are used in the same model.

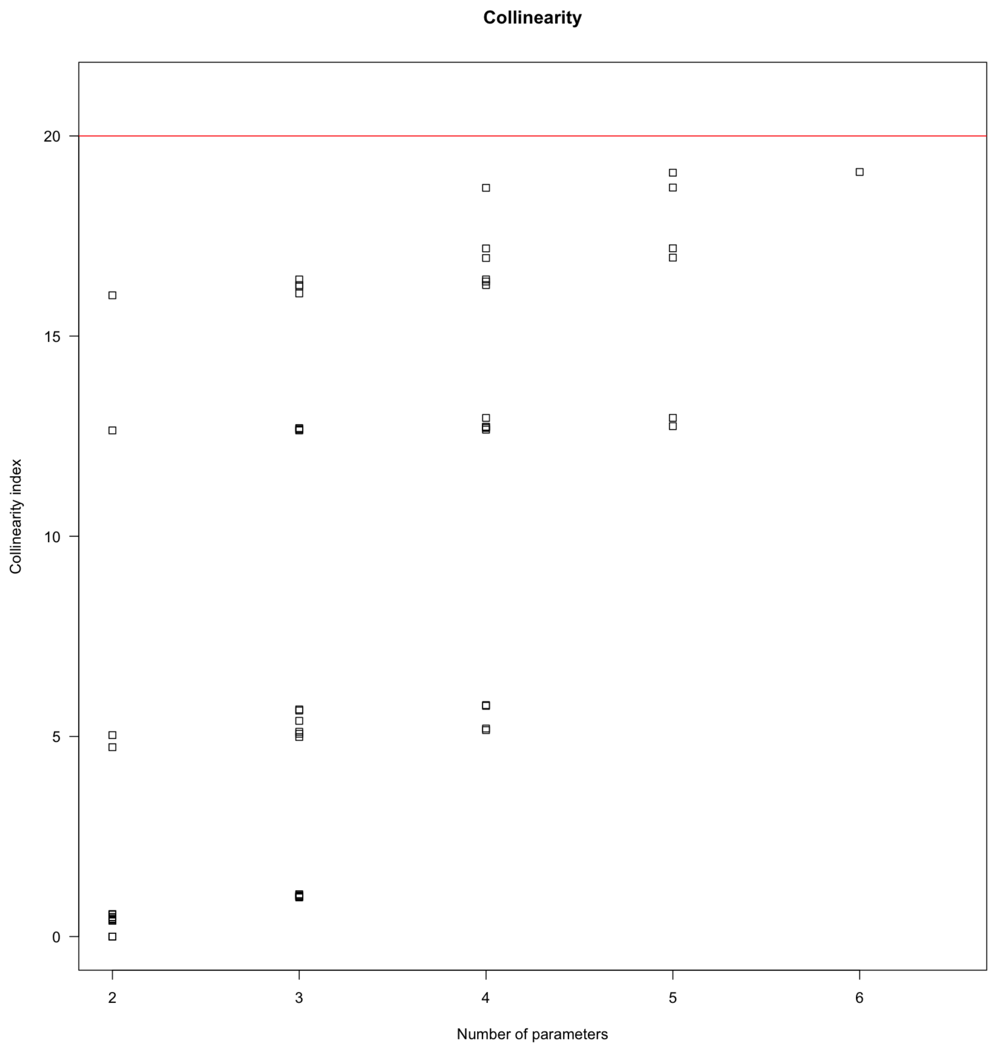

Figure 7: Collinearity analysis

The parameter set that yielded best fit was used as initial parameter values for running a Markov chain Monte Carlo (MCMC)[8] using the function ’modMCMC’ based on the delayed rejection adaptive Metropolis algorithm. As the model has several variables (observed datapoints) with different magnitudes, it is better to scale each variable independently. Hence, the initial model variance for MCMC was assumed to be the ’mean of the unweighted squared residuals’ as returned from ’modFit’. MCMC was run for 10000 iterations and after every 10 iterations, the parameter covariance matrix was (re)evaluated based on the parameters kept thus far, and was used to update the proposal distribution.

### 8 Main model fit

The model could accurately reproduce the number of HIV-diagnosed MSM in Switzerland and the syphilis cases among the MSM with and without HIV diagnosis. (Manuscript figure 2) In addition, the model provided an estimate for incidence rate of syphilis among the MSM with and without HIV diagnosis stratified by reported non-steady partners (Manuscript figure 3).

Posterior distribution of the fitted parameters are given in table 1.
Figure 8 shows the trace of parameter values (jump) of an MCMC chain. Corresponding sum of squares of the MCMC chain is shown in Figure 9. The plotted results demonstrate (near-)convergence of the chain. Overall 62% of the iterations were accepted (6242 of 10000 iterations). The pairwise sensitivity among the fitted parameters were assessed by calculating the correlation using 'pairs' function as shown in Figure 10. Figure 11 shows the posterior distribution of the fitted parameters based on the *ad-hoc* prior bounds.

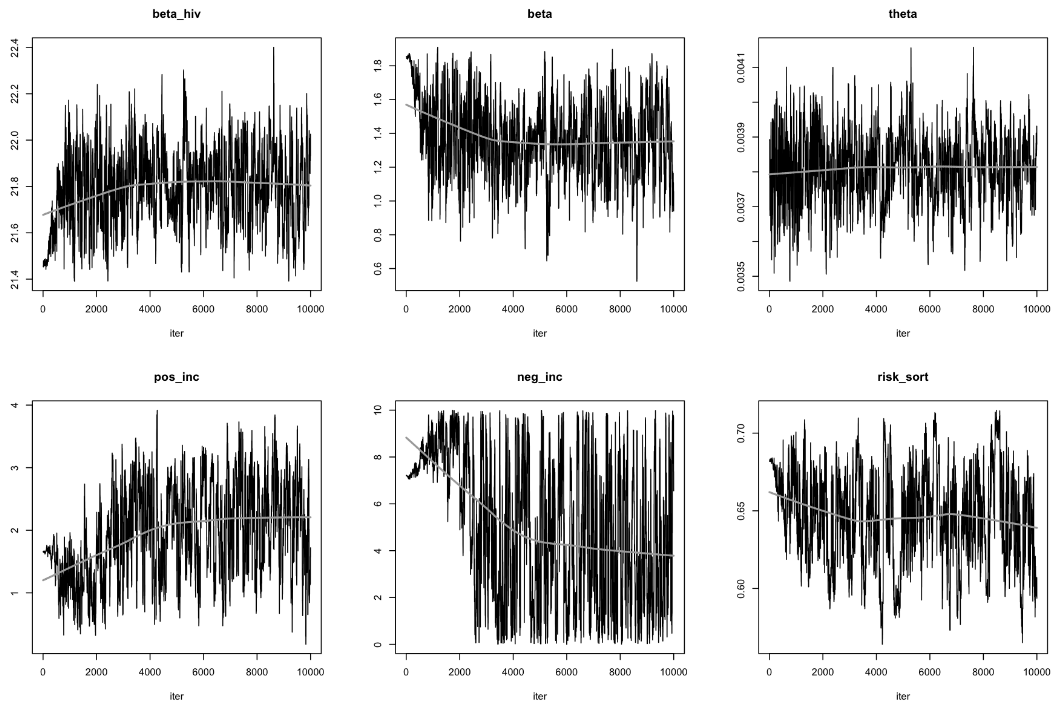

Figure 8: Trace of parameter values of an MCMC chain

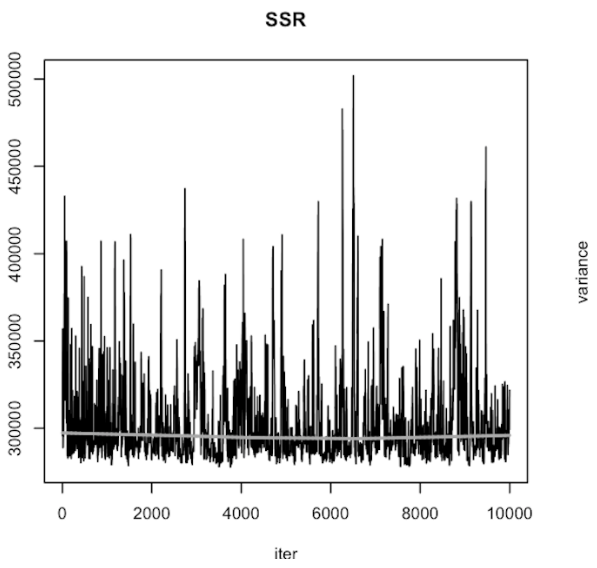

Figure 9: Sum of squares for each iteration of MCMC chain

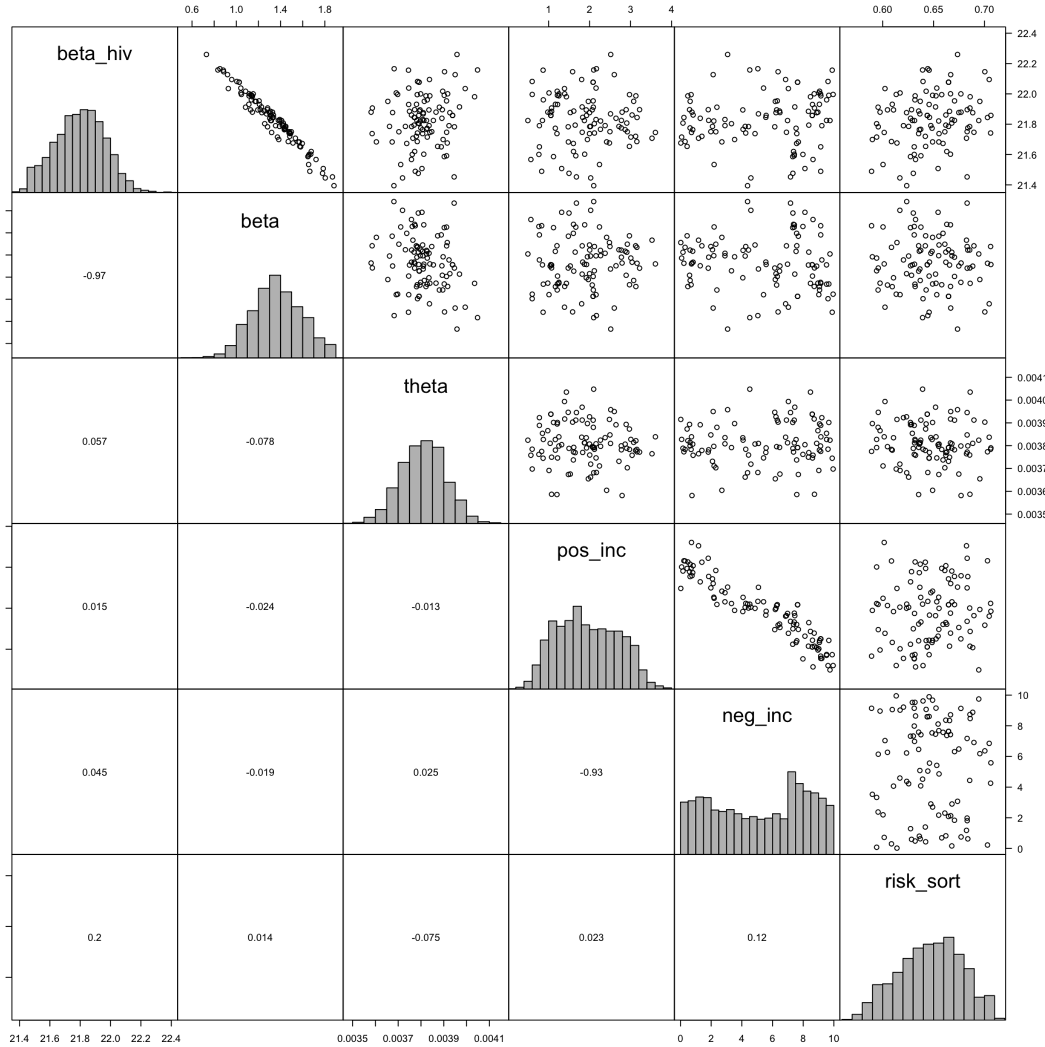

Figure 10: Pairs plot of MCMC results

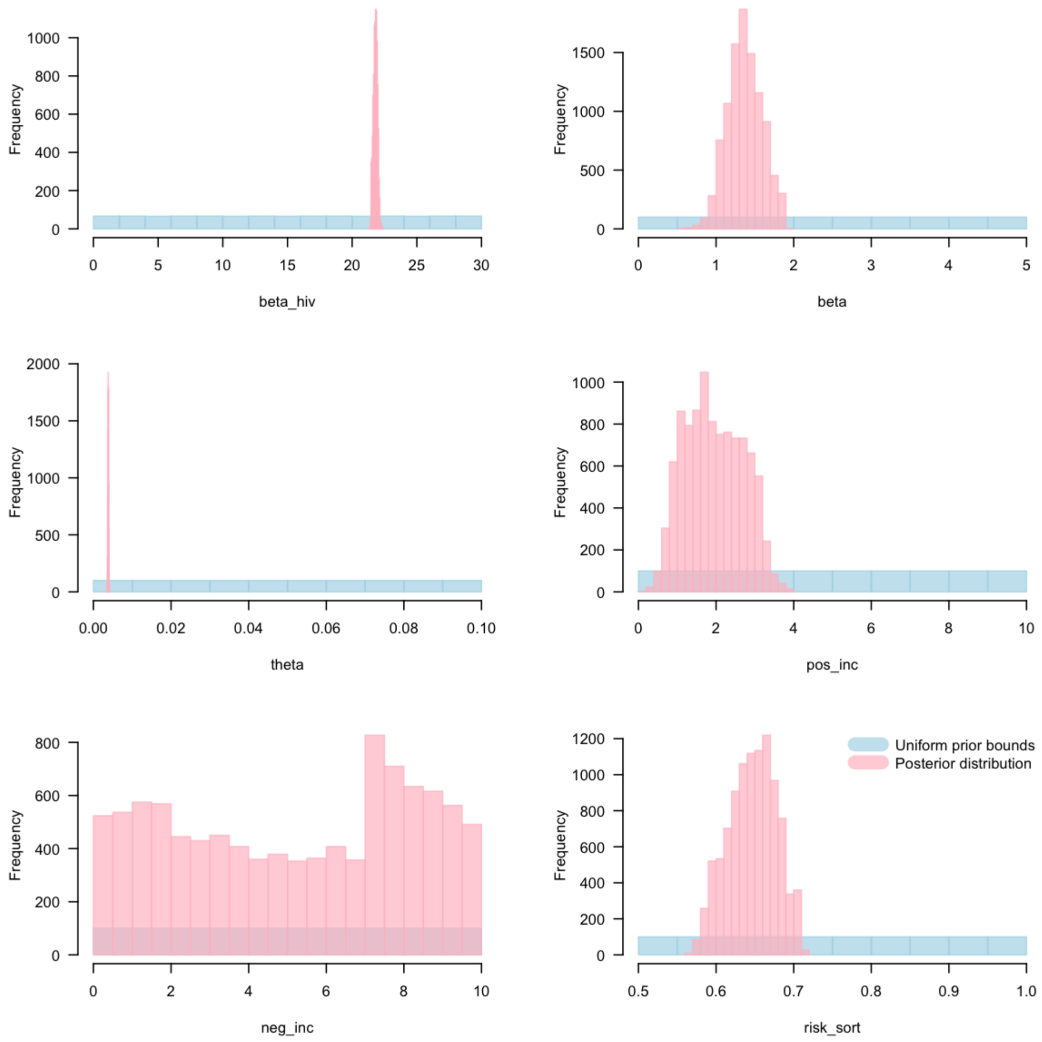

Figure 11: Prior and posterior distribution of free parameters

We further ran two more independent chains of MCMC and used the package 'coda' to perform the diagnostic tests of convergence to the equilibrium distribution of the Markov chains[9]. We performed Gelman and Rubin's convergence diagnostics on MCMC chains using the function 'gelman.diag'. We estimated the 'potential scale reduction factor' for each variable in MCMC chain along with the upper 95% confidence interval as shown in table . As the upper CI is close to 1, approximate convergence is diagnosed. For multivariate chains, a multivariate value was calculated that bounds above the potential scale reduction factor for any linear combination of the parameters and found the value to be 1.07. This indicates the convergence of chains.

Table 3: Potential scale reduction

| Variable | Point estimate | Upper CI |
|---|---|---|
| $\beta_0$ | 1.01 | 1.01 |
| $\beta_1$ | 1.00 | 1.01 |
| $\theta$ | 1.02 | 1.07 |

| | | |
|---|---|---|
| $f$ | 1.01 | 1.04 |
| $\inf_0(0)$ | 1.02 | 1.06 |
| $\inf_1(0)$ | 1.04 | 1.14 |

### 9 Counterfactual scenarios

We compared incidence of syphilis obtained with the main model fit with that simulated when assuming alternative scenarios.

We considered the impact of change in transmission risk between the years 2006 and 2018. With no change in the proportion of MSM switching from MSM with nsP to MSM without nsP and vice-versa between the years 2006 and 2018, , we estimated a reduction of 17.56% (4.04 vs 4.91 cases per 1000 person-years) in incidence rate of syphilis in 2017 (Figure 12)

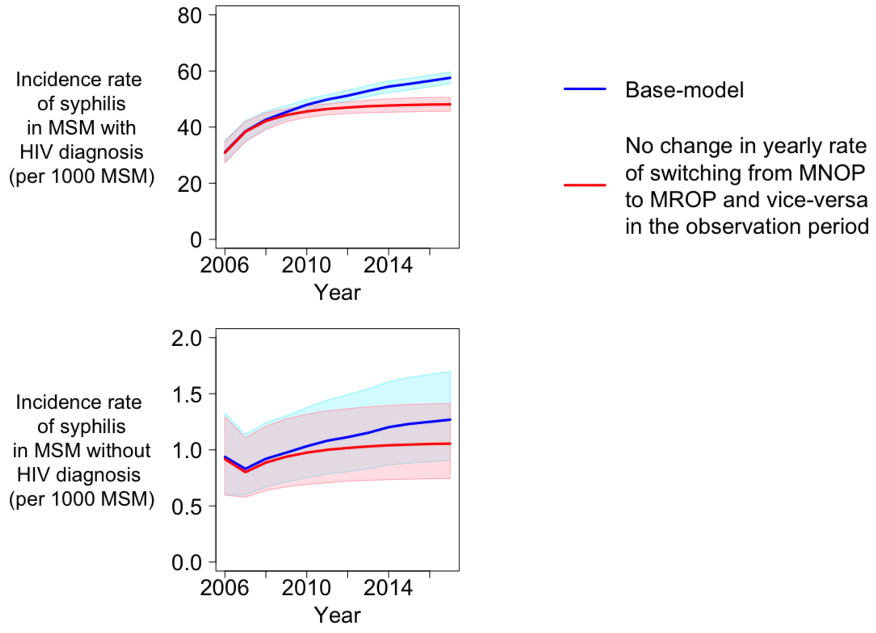

Figure 12: Counterfactual scenario: impact of change in transmission risk

### 10 Sensitivity analysis

To test the robustness of the model and possible inconsistencies in data, we recalibrated the model and determined the corresponding change in the incidence rate of syphilis, the fitted parameters and the goodness of fit (sum of squared weighted residuals). Firstly, we considered alternative transmission risk. We assumed MSM reporting condomless anal intercourse as transmission risk instead of MSM with non-steady partners. (Figure 13)

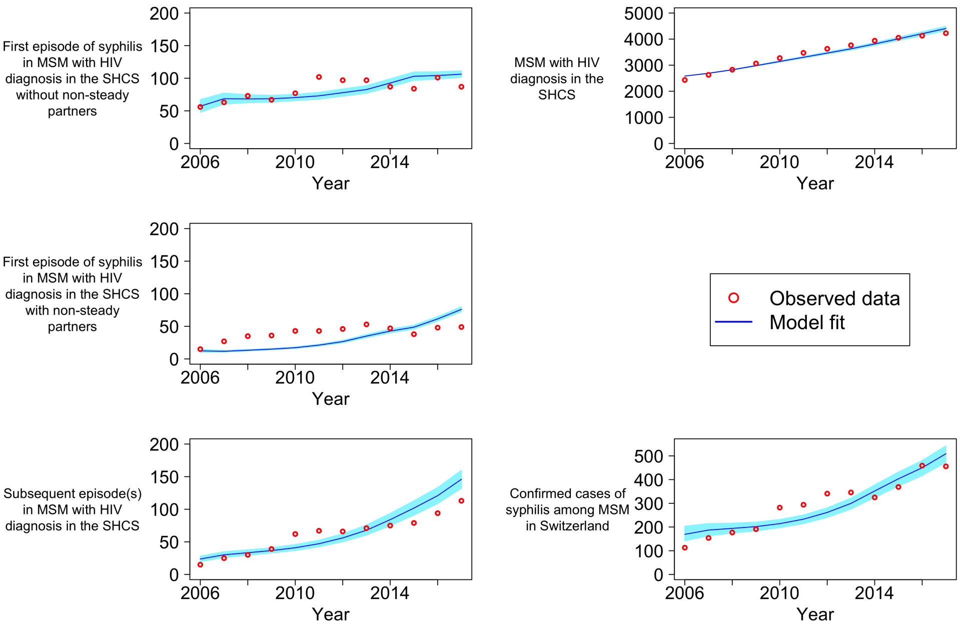

Figure 13: Sensitivity analysis: Model fit with MSM reporting condomless anal intercourse as transmission risk of syphilis

Secondly, we assessed the impact of an alternative assumption for the transmission rate of syphilis among MSM with and without HIV diagnosis. There was an apparent contradiction in the data, despite similar transmission risk of syphilis among MSM with and without HIV diagnosis, we observed a higher incidence of syphilis among MSM with HIV diagnosis than those without HIV diagnosis even after adjusting for contact probabilities. To resolve this contradiction, in the main model, we assumed different transmission rates of syphilis in MSM with and without HIV diagnosis in our model. In the absence of this assumption, the model is unable to reproduce the syphilis epidemic in Switzerland and led to either a strong overestimation of syphilis incidence in MSM without HIV diagnosis or a strong underestimation in MSM with HIV diagnosis. (Figure 14)

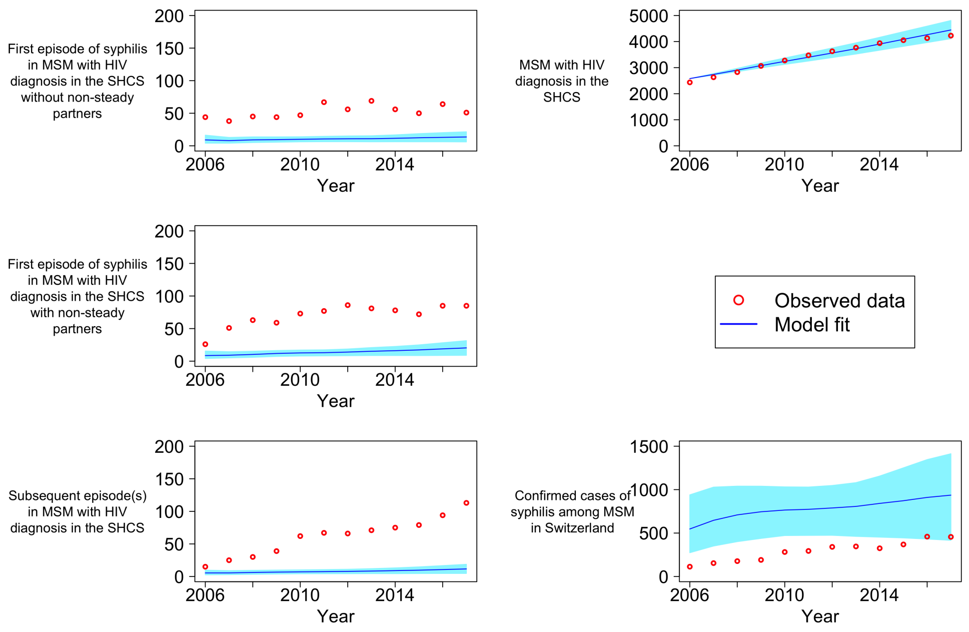

Figure 14: Sensitivity analysis: Model fit assuming no different transmission rates of syphilis in MSM with and without HIV diagnosis

Alternatively, we assumed that there is no difference in transmission rate of syphilis among MSM with and without HIV diagnosis and fitted a parameter to account for possible underreporting of syphilis cases to FOPH among MSM without HIV diagnosis. (Figure 15)

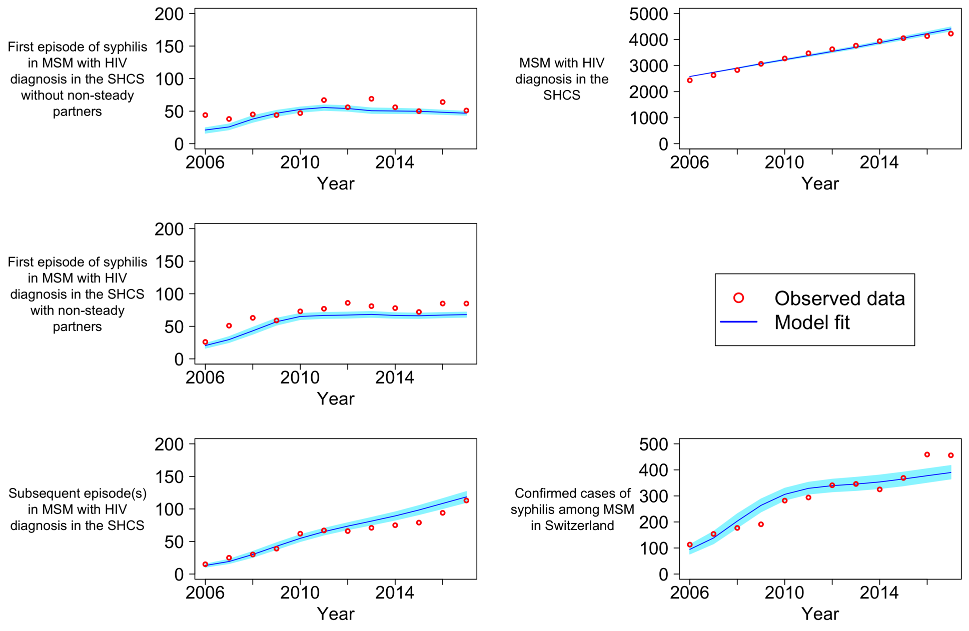

Figure 15: Sensitivity analysis: Model fit assuming no different transmission rates of syphilis in MSM with and without HIV diagnosis and a fitted parameter to account for possible underreporting of syphilis cases to FOPH among MSM without HIV diagnosis

Thirdly, we investigated the impact of a possible overestimation of transmission risk among MSM without HIV diagnosis by assuming the proportion of MSM without HIV diagnosis and with nsP to be 25%, 50%

and 75% lower than the proportion of MSM without HIV diagnosis and with nsP estimated from data published in the Gay survey (Figure 16)

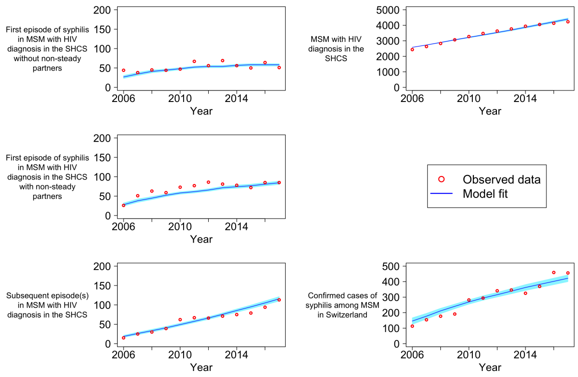

(a) 25% lower proportion of MSM without HIV diagnosis and with nsP

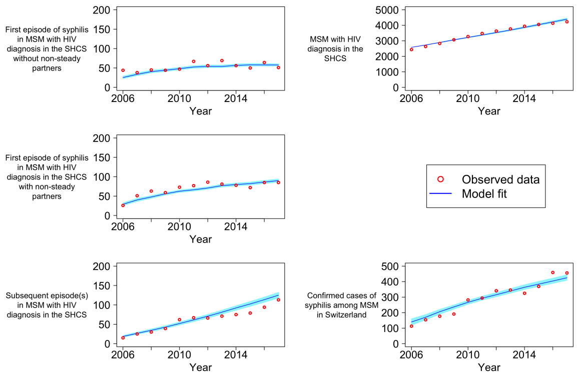

(b) 50% lower proportion of MSM without HIV diagnosis and with nsP

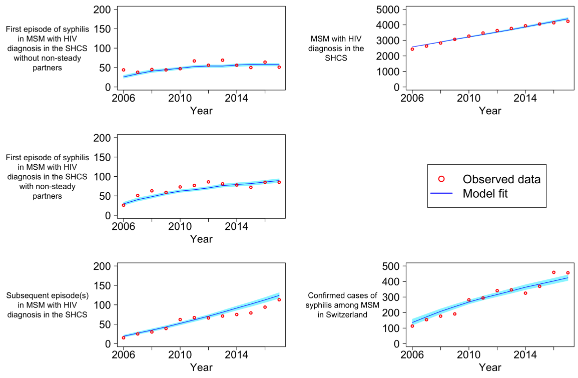

(c) 75% lower proportion of MSM without HIV diagnosis and with nsP

Figure 16: Sensitivity analysis: Model fit assuming the proportion of MSM without HIV diagnosis and with nsP to be 25%, 50% and 75% lower than the proportion of MSM without HIV diagnosis and with nsP estimated from data published in the Gay survey

Fourthly, we assessed the impact of infectiousness of syphilis in latent stage on dependency between the screening frequency and the syphilis incidence by assuming 1% and 10% infectiousness during the latent stage of syphilis (>3 months) of that in the primary and secondary stages of syphilis (<3 months) instead of 0% as assumed in the main model fit. (Figure 17) By means of counterfactual scenarios, we assessed the impact of infectiousness during the latent stage of syphilis on the reduction in syphilis incidence in 2017 when the screening frequency for syphilis among MSM with HIV diagnosis and with nsP was increased from once per year to twice per year during the observation period and found the reduction in syphilis incidence to decrease with increase in infectiousness during the latent stage (Manuscript figure 5)

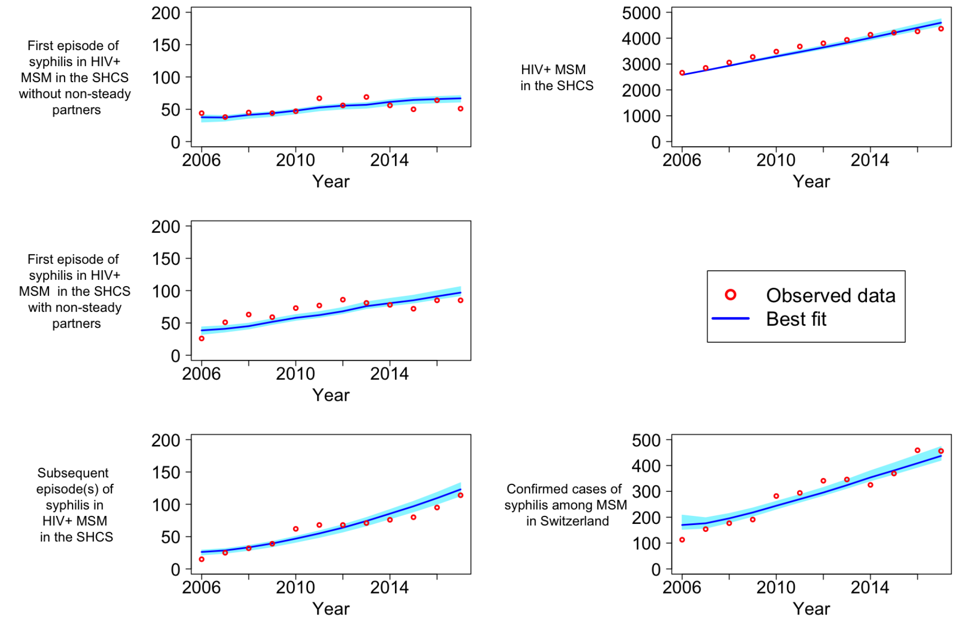

(a) 1% infectiousness during the latent stage of syphilis

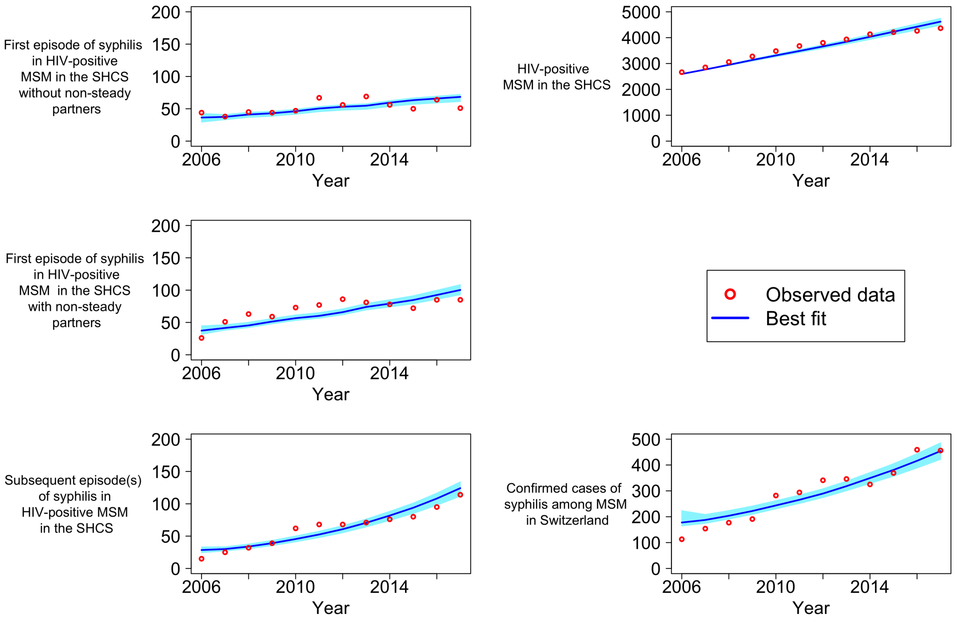

(b) 10% infectiousness during the latent stage of syphilis

Figure 17: Sensitivity analysis: Model fit assuming the infectiousness during the latent stage of syphilis to be 1% and 10% of that in the primary and secondary stages of syphilis.

Finally, we tested the effect of using an alternative fitting procedure. Instead of fitting the model through minimization of the sum of squared weighted residuals, we fitted the model through maximization of the likelihood by assuming Poisson distributed incident cases of syphilis. (Figure 18)

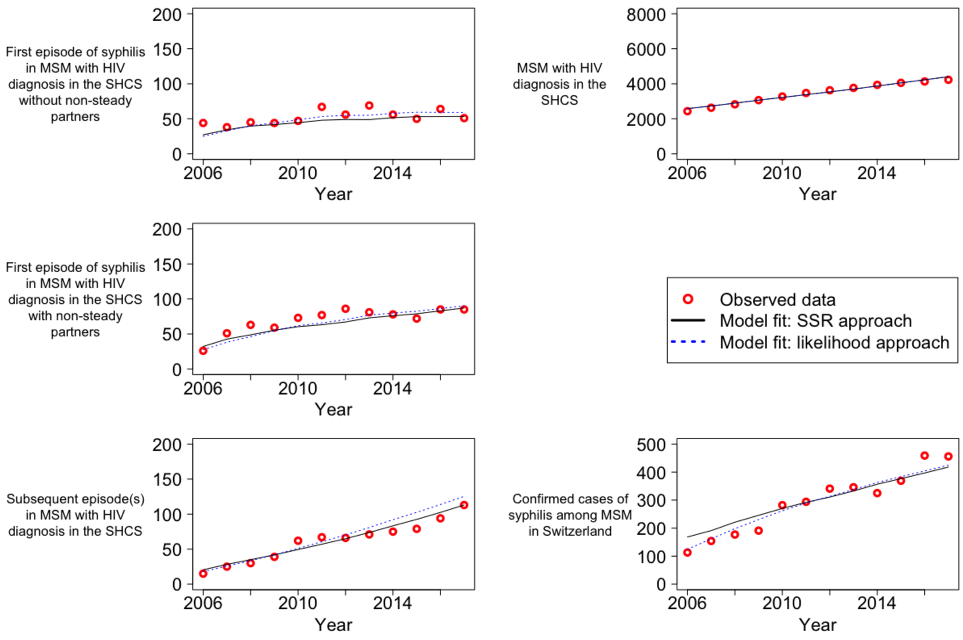

Figure 18: Sensitivity analysis: Comparing model fitting procedures

Table 4: Fitted parameters for main model fit and re-calibrated models (sensitivity analysis)

| Model description | $\beta_0$ ($year^{-1}$) | $\beta_1$ ($year^{-1}$) | $\theta$ ($year^{-1}$) | $f$ | $inf_0(0)$ | $inf_1(0)$ | Goodness of fit (SSR) | Median IR (per 1000 py) | Model fit |
|---|---|---|---|---|---|---|---|---|---|
| Main model | 1.27 | 21.88 | 0.0038 | 0.64 | 0.38 | 2.97 | 148.3 | 3.77 | Manuscript figure 2 |
| MSM with nsCAI as transmission risk of syphilis instead of MSM with nsP | 1.12 | 21.11 | 0.0060 | 0.64 | 0.16 | 4.51 | 413.0 | 3.10 | Figure 13 |
| Same transmission rates of syphilis in MSM with and without HIV diagnosis | 7.41 * | - * | 0.00450 | 0.50 | 2.22 | 2.27 | 12526.1 | 9.77 | Figure 14 |
| Same transmission rates of syphilis in MSM with and without HIV diagnosis and fitted a parameter to account for possible underreporting of syphilis cases to FOPH | 7.64 † | - † | 0.0038 | 0.57 | 0.73 | 9.71 | 206.6 | 4.19 | Figure 15 |
| Proportion of MSM without HIV diagnosis and with nsP to be 25% lower than that used in the main model | 1.51 | 21.62 | 0.0051 | 0.59 | 0.73 | 2.62 | 136.4 | 3.82 | Figure 16 (a) |
| Proportion of MSM without HIV diagnosis and with nsP to be 50% lower than that used in the main model | 1.31 | 21.86 | 0.0076 | 0.62 | 1.25 | 2.40 | 124.4 | 3.81 | Figure 16 (b) |
| Proportion of MSM without HIV diagnosis and with nsP to be 75% lower than that used in the main model | 1.43 | 21.73 | 0.0155 | 0.60 | 0.13 | 2.68 | 123.6 | 3.81 | Figure 16 (c) |
| Infectiousness during latent stage of syphilis to 1% of that in primary and secondary stage of syphilis | 1.04 | 21.15 | 0.0038 | 0.58 | 0.27 | 4.50 | 154.27 | 3.59 | Figure 17 (a) |

| | | | | | | | | | |
|---|---|---|---|---|---|---|---|---|---|
| Infectiousness during latent stage of syphilis to 10% of that in primary and secondary stage of syphilis | 0.74 | 16.17 | 0.0037 | 0.54 | 9.96 | 9.97 | 397.6 | 2.98 | Figure 17 (b) |
| Model fitting through maximization of the likelihood by assuming Poisson distributed incident cases of syphilis | 1.09 | 22.02 | 0.0040 | 0.63 | 0.03 | 2.58 | - ** | 3.77 | Figure 18 |

* We assumed $\beta_1 = \beta_0$ and we used an ad-hoc prior bound of (0-30) year$^{-1}$ for $\beta_0$

† We assumed $\beta_1 = \beta_0$ and we used an ad-hoc prior bound of (0-30) year$^{-1}$ for $\beta_0$. We further assumed an ad-hoc prior bound of (1-$\infty$) for the parameter ($\zeta$) to account for possible underreporting of syphilis cases to FOPH and estimated $\zeta$ to be 40.84. *i.e.*, incidence of syphilis among MSM without HIV diagnosis is 45.57 times the incidence of syphilis in MSM without HIV diagnosis reported to FOPH.

** The model fitting was done by maximization of the likelihood and not by minimizing SSR.

[0]SSR = Sum of squared weighted residuals; IR = incidence rate; py = person-years; nsCAI = condomless anal intercourse with non-steady partners; nsP = non-steady partners; FOPH = The Federal Office for Public Health;
